# Preclinical interstitial lung disease in relatives of familial pulmonary fibrosis patients

**DOI:** 10.1101/2022.04.27.22274348

**Authors:** Sionne EM Lucas, Kelsie Raspin, John Mackintosh, Ian Glaspole, Paul N Reynolds, Collin Chia, Christopher Grainge, Peter Kendall, Lauren Troy, David A Schwartz, Richard Wood-Baker, Simon LF Walsh, Yuben Moodley, James Robertson, Sacha Macansh, Eugene H Walters, Daniel Chambers, Tamera J Corte, Joanne L Dickinson

**Affiliations:** Menzies Institute for Medical Research, University of Tasmania, Hobart, TAS, Australia; School of Medicine, The University of Queensland, Brisbane, QLD, Australia; QLD Lung Transplant Service, Department of Thoracic Medicine, The Prince Charles Hospital, Brisbane, QLD, Australia; Department of Respiratory Medicine, Alfred Health, Melbourne, VIC, Australia; Department of Medicine, Monash University, Melbourne, VIC, Australia; Royal Adelaide Hospital, Adelaide, SA, Australia; University of Adelaide, Adelaide, SA, Australia; Launceston General Hospital, Launceston, TAS, Australia; School of Medicine and Public Health, University of Newcastle, Callaghan, NSW, Australia; Respiratory Medicine Service, Albany, WA, Australia; School of Medicine and Pharmacology, University of Western Australia, Perth, WA, Australia; Department of Respiratory Medicine, Royal Prince Alfred Hospital, Camperdown, NSW, Australia; School of Medicine, The University of Sydney, Camperdown, NSW, Australia; Department of Medicine, University of Colorado Anschutz Medical Campus, Aurora, CO, USA; School of Medicine, University of Tasmania, Hobart, TAS, Australia; National Heart and Lung Institute, Imperial College London, London, England, UK; University of Western Australia, Institute for Respiratory Health, Perth, WA, Australia; Department of Respiratory Medicine, Fiona Stanley Hospital, Perth, WA, Australia; Border Physicians Group, West Albury, NSW, Australia; Lung Foundation Australia, Brisbane, QLD, Australia

## Abstract

Family history is amongst the strongest risk factors for interstitial lung disease (ILD), with emerging evidence for a shared genetic aetiology across ILD subtypes. Recruited families comprised at least two first-degree relatives who had been previously diagnosed with an ILD. All living cases and available unaffected first-degree relatives underwent a clinical examination for evidence of ILD. Preclinical ILD was diagnosed in 47.7% of first-degree relatives who had previously self-reported as unaffected. This study highlights the strong genetic predisposition in family members of ILD cases, and supports the call for routine screening of individuals with a family history of ILD.

---

Interstitial lung disease (ILD) is a group of heterogeneous disorders characterised by chronic lung inflammation and/or fibrosis. Idiopathic pulmonary fibrosis (IPF) is one of the most common and devastating forms of ILD and is frequently diagnosed at an advanced stage, by which time patients have a life-expectancy of 2-3 years. There is growing evidence that genetic factors involved in IPF also contribute to a broader range of ILDs and interstitial lung abnormalities (ILA)^1^. ILA is defined as incidental findings of specific computed tomography features that affect more than 5% of any lung zone and are potentially compatible with ILD (i.e. ground glass or reticular abnormalities, traction bronchiectasis, honeycombing and non-emphysematous cysts)^2^. A growing number of studies have reported evidence of ILAs in clinically unaffected relatives of IPF patients^3-5^. Indeed, the recent Fleischner Society’s position paper recommends that the identification of ILA in familial ILD should not be considered incidental and suggests use of the term “preclinical ILD” ^2^, and this terminology has been adopted here. Our study has established a clinically-annotated familial ILD genetic resource aiming to better understand disease causation. Recently, studies have suggested that screening first-degree relatives of ILD and IPF patients for evidence of preclinical ILD may be worthwhile approach to improve detection of ILD in its early stages^3-5^. Here, we sought to provide additional evidence through sharing insights generated from a familial ILD cohort.

## METHOD

This study defined familial ILD as a family with two or more first degree relatives that had previously been diagnosed with ILD or ILA (hereafter collectively termed “patients”), where at least one had IPF. Patients with familial ILD were identified via a questionnaire sent to all living Australian IPF Registry^6^ participants (September to November 2019) or directly recruited by participating respiratory physicians. Index patients were interviewed to determine the eligibility of the family, and if suitable, all living patients and at least one self-reported unaffected first-degree relative (hereafter known as “relative”) were invited to participate. Informed consent and a physical examination were conducted by respiratory physicians, after which participants completed: questionnaires, including the St. George’s Respiratory Questionnaire (SGRQ)^7^; a chest high resolution computed tomograph (HRCT) scan; formal pulmonary function testing (PFT); and blood/saliva collection. Pre-transplant PFT and HRCT scans were sourced for patients who had received a lung transplant. HRCT images were blinded and assessed by an ILD physician (JM) and preclinical ILD was classified using the Fleischner Society’s ILA definition^2^. This report includes all families in which at least one unaffected relative was screened.

## RESULTS

Fifteen relatives from 12 families were screened (for example, **Figure 1**) including ten siblings, three children of patients, and two individuals who had both an affected parent and sibling. Seven relatives were found to have preclinical ILD (46.7%), while eight (53.3%) had no evidence of abnormalities at the time of examination. Notably, only siblings of patients were found to have preclinical ILD, with a prevalence of 58.3% when considering relatives with an affected sibling.

**Figure 1:**
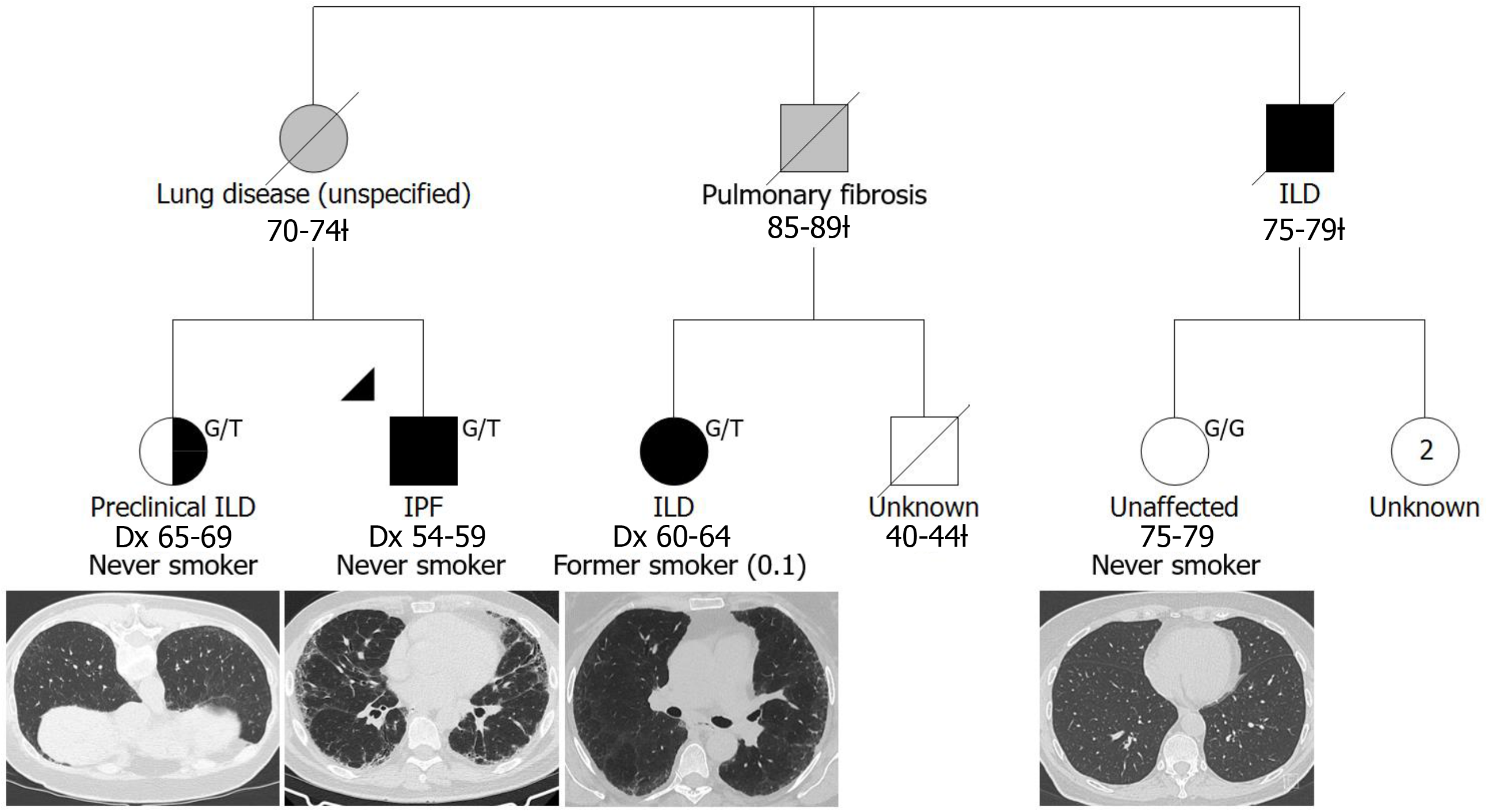
A simplified pedigree depicting a family in which two relatives were screened: one was diagnosed with preclinical ILD while the other was unaffected at the time of examination. Males and females are depicted by squares and circles (respectively); deceased individuals are indicated by a diagonal slash and the age range at death is indicated below, designated by a □ clinical diagnosis is as reported by the living generation; age range at examination for the unaffected relative is indicated below; ‘Dx’ indicates age range at diagnosis; two living, unscreened female relatives are currently 64-69 years of age; self-reported smoking status is indicated and pack years is presented in brackets; *MUC5B* rs35705950 genotype is presented at the top right, G/T indicates a heterozygous carrier and G/G indicates homozygous wildtype; representative chest HRCT images are presented below study participants.

Relatives diagnosed with preclinical ILD were older (71.6 *vs* 60.8 years), more likely to be female and had lower body mass index (BMI) than unaffected relatives **(Table 1)**. Rates of ever smoking was higher in known patients (54.5%) and unaffected relatives (50.0%), compared to the preclinical ILD group (28.6%). Examination of the established IPF risk variant in the *MUC5B* promotor (rs35705950) revealed that unaffected relatives were less likely to be carriers of the risk allele (‘T’), with a frequency of 18.8% compared to 45.2% in the known patients and 35.7% in the relatives with preclinical ILD.

**Table 1.**
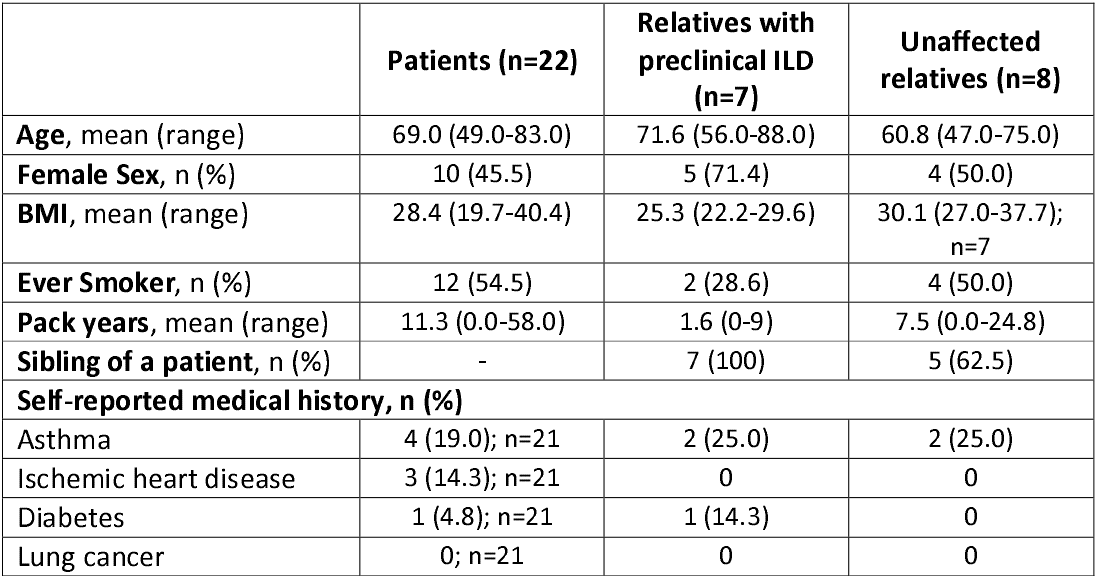
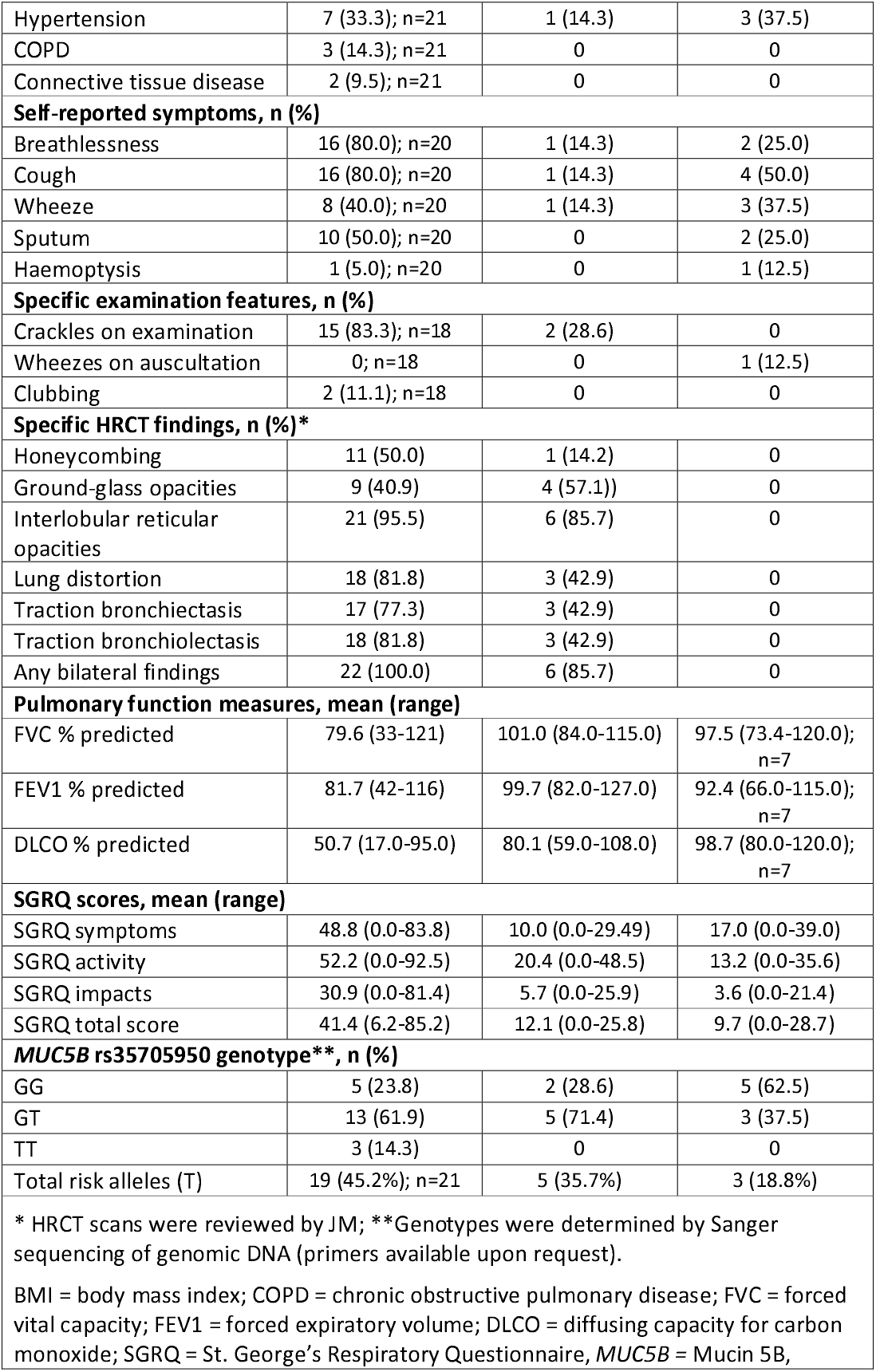
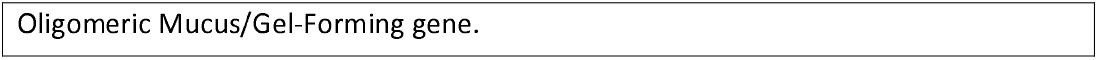
Clinical characteristics of participants at the time of recruitment.

|  | **Patients (n=22)** | **Relatives with preclinical ILD (n=7)** | **Unaffected relatives (n=8)** |
| --- | --- | --- | --- |
| **Age**, mean (range) | 69.0 (49.0-83.0) | 71.6 (56.0-88.0) | 60.8 (47.0-75.0) |
| **Female Sex**, n (%) | 10 (45.5) | 5 (71.4) | 4 (50.0) |
| **BMI**, mean (range) | 28.4 (19.7-40.4) | 25.3 (22.2-29.6) | 30.1 (27.0-37.7);<br>n=7 |
| **Ever Smoker**, n (%) | 12 (54.5) | 2 (28.6) | 4 (50.0) |
| **Pack years**, mean (range) | 11.3 (0.0-58.0) | 1.6 (0-9) | 7.5 (0.0-24.8) |
| **Sibling of a patient**, n (%) | - | 7 (100) | 5 (62.5) |
| **Self-reported medical history, n (%)** |  |  |  |
| Asthma | 4 (19.0); n=21 | 2 (25.0) | 2 (25.0) |
| Ischemic heart disease | 3 (14.3); n=21 | 0 | 0 |
| Diabetes | 1 (4.8); n=21 | 1 (14.3) | 0 |
| Lung cancer | 0; n=21 | 0 | 0 |
| Hypertension | 7 (33.3); n=21 | 1 (14.3) | 3 (37.5) |
| COPD | 3 (14.3); n=21 | 0 | 0 |
| Connective tissue disease | 2 (9.5); n=21 | 0 | 0 |
| **Self-reported symptoms, n (%)** |  |  |  |
| Breathlessness | 16 (80.0); n=20 | 1 (14.3) | 2 (25.0) |
| Cough | 16 (80.0); n=20 | 1 (14.3) | 4 (50.0) |
| Wheeze | 8 (40.0); n=20 | 1 (14.3) | 3 (37.5) |
| Sputum | 10 (50.0); n=20 | 0 | 2 (25.0) |
| Haemoptysis | 1 (5.0); n=20 | 0 | 1 (12.5) |
| **Specific examination features, n (%)** |  |  |  |
| Crackles on examination | 15 (83.3); n=18 | 2 (28.6) | 0 |
| Wheezes on auscultation | 0; n=18 | 0 | 1 (12.5) |
| Clubbing | 2 (11.1); n=18 | 0 | 0 |
| **Specific HRCT findings, n (%)\*** |  |  |  |
| Honeycombing | 11 (50.0) | 1 (14.2) | 0 |
| Ground-glass opacities | 9 (40.9) | 4 (57.1)) | 0 |
| Interlobular reticular opacities | 21 (95.5) | 6 (85.7) | 0 |
| Lung distortion | 18 (81.8) | 3 (42.9) | 0 |
| Traction bronchiectasis | 17 (77.3) | 3 (42.9) | 0 |
| Traction bronchiolectasis | 18 (81.8) | 3 (42.9) | 0 |
| Any bilateral findings | 22 (100.0) | 6 (85.7) | 0 |
| **Pulmonary function measures, mean (range)** |  |  |  |
| FVC % predicted | 79.6 (33-121) | 101.0 (84.0-115.0) | 97.5 (73.4-120.0); n=7 |
| FEV1 % predicted | 81.7 (42-116) | 99.7 (82.0-127.0) | 92.4 (66.0-115.0); n=7 |
| DLCO % predicted | 50.7 (17.0-95.0) | 80.1 (59.0-108.0) | 98.7 (80.0-120.0); n=7 |
| **SGRQ scores, mean (range)** |  |  |  |
| SGRQ symptoms | 48.8 (0.0-83.8) | 10.0 (0.0-29.49) | 17.0 (0.0-39.0) |
| SGRQ activity | 52.2 (0.0-92.5) | 20.4 (0.0-48.5) | 13.2 (0.0-35.6) |
| SGRQ impacts | 30.9 (0.0-81.4) | 5.7 (0.0-25.9) | 3.6 (0.0-21.4) |
| SGRQ total score | 41.4 (6.2-85.2) | 12.1 (0.0-25.8) | 9.7 (0.0-28.7) |
| ***MUC5B* rs35705950 genotype\*\*, n (%)** |  |  |  |
| GG | 5 (23.8) | 2 (28.6) | 5 (62.5) |
| GT | 13 (61.9) | 5 (71.4) | 3 (37.5) |
| TT | 3 (14.3) | 0 | 0 |
| Total risk alleles (T) | 19 (45.2%); n=21 | 5 (35.7%) | 3 (18.8%) |
| * HRCT scans were reviewed by JM; **Genotypes were determined by Sanger sequencing of genomic DNA (primers available upon request).<br>BMI = body mass index; COPD = chronic obstructive pulmonary disease; FVC = forced vital capacity; FEV1 = forced expiratory volume; DLCO = diffusing capacity for carbon monoxide; SGRQ = St. George's Respiratory Questionnaire, *MUC5B* = Mucin 5B, |  |  |  |

Oligomeric Mucus/Gel-Forming gene.

## DISCUSSION

We observed a high incidence of preclinical ILD in relatives of patients with familial ILD in our Australian cohort (46.7%), notably higher than recent studies in the American families described in Hunninghake et al.^3^ (26%) and Salisbury et al.^5^ (23%). It is possible that the incidence of preclinical ILD was higher in our cohort due to our targeted selection of families with a strong family history of ILD, although our study was also smaller and older (65.8 years compared to 60 years^3^ and 53.1 years^5^), which may have contributed to the differences. Preclinical ILD was only observed in siblings of patients, consistent with Aburto et al.^4^ who demonstrated that 52.6% of siblings of apparent sporadic IPF patients had preclinical ILD, whilst only 5.6% of patient’s offspring were affected at the time of examination. The mean age of the offspring cohort in Aburto et al.^4^ was 47.7 years and, as the mean age of familial IPF onset is 55 years^8^, some individuals may be yet to develop preclinical ILD. Neither Hunninghake et al.^3^ or Salisbury et al.^5^ differentiated between siblings and offspring. Further research is required to determine an appropriate screening window for the offspring of ILD patients, however, we propose that the age of diagnosis for known patients in the family is an important consideration.

The *MUC5B* risk allele was nearly twice as frequent in relatives with preclinical ILD than in unaffected relatives (35.7% *vs* 18.8%) and almost as frequent as observed in known patients (45.2%), consistent with the findings of Hunninghake et al.^3^ and Salisbury et al.^5^ and highlights the strong genetic predisposition in our familial cohort. We observed a lower rate of smoking (both ever smoking and pack years) in the relatives found to have preclinical ILD compared to their unaffected counterparts, however the preclinical ILD group were more likely to be female and on average ten years older, and the group sizes were small. This finding, however, is consistent with Hunninghake et al.^3^ who reported a lower frequency of ever smoking in relatives from families with familial ILD (28%) compared to families with apparent sporadic disease (52%).

Our study provides further evidence that clinical examination of first-degree relatives of ILD patients, particularly those with a strong family history, facilitates early diagnosisis, which critical to providing individuals with opportunities to adopt healthy lifestyle interventions and access to innovative clinical management practices as they are developed.

## Data Availability

All data produced in the present study are available upon reasonable request to the authors

## ETHICS APPROVAL

This study was conducted in accordance with the amended Declaration of Helsinki. The study was approved by the Sydney Local Health District Human Research Ethics Committee (X18-0193), University of Tasmania Health and Medical Human Research Ethics Committee (17775) and the Prince Charles Hospital Human Research Ethics Committee (53369).

## ACKNOWLEDGEMENTS

We are greatly indebted to our participants and their families for volunteering their time and clinical data; without their participation, this study would not be possible. We also thank the ILD nurses and coordinators, including Catriona Doran from Royal Adelaide Hospital; Elizabeth Ray, Shannon Cleary, Jessica Rhodes and Qi Lin from Royal Prince Alfred Hospital; Amy Cashmore from John Hunter Hospital; Sandra Bancroft from The Prince Charles Hospital; Karen Symons from The Alfred; Shona Knights-Rennie from Border Physicians Group; Samantha Wadham and Helen Rodgers from Sunshine Coast University Hospital and; David Clark from PathWest Albany. The Australian IPF Registry is facilitated by Lung Foundation Australia and is supported by unrestricted educational grants from Foundation Partners, Boehringer Ingelheim and Roche Products, Pty. Limited. SEML, PR, LT, DAS, RWB, SW, YM, SM, HW, DC, TJC and JLD made substantial contributions to the conception and design of the study; SL, KR, SM, TJC, LT, IG, PNR, DC, CC, CG, JR and PK made substantial contributions to the acquisition of patients and clinical data; SEML and JM conducted data analysis; SEML; KR; JM and TLC and JLD made substantial ontributions to the interpretation of the data; SEML and JLD drafted the manuscript; while all authors were involved in revising the manuscript critically for important intellectual content; gave final approval for publishing; and agreed to be accountable for all aspects of the work in ensuring that questions related to the accuracy or integrity of any part of the work are appropriately investigated and resolved. SEML, TJC and JLD are guarantors of this work, taking responsibility for the integrity of the work as a whole, from incepton to published article.

